# Evaluation of real and perceived risk to health care workers caring for patients with the Omicron variant of the SARS-CoV-2 virus in surgery and obstetrics

**DOI:** 10.1101/2022.10.30.22281627

**Authors:** Chaithanya Nair, Robert Kozak, Nasrin Alavi, Hamza Mbareche, Rose C. Kung, Kellie E. Murphy, Darian Perruzza, Stephanie Jarvi, Elsa Salvant, Noor Niyar N. Ladhani, Albert J.M. Yee, Louise-Helene Gagnon, Richard Jenkinson, Grace Y. Liu, Patricia E. Lee

**Affiliations:** Temerty Faculty of Medicine, University of Toronto, Toronto, Ontario, Canada; Division of Microbiology, Department of Medicine, Sunnybrook Health Sciences Centre, University of Toronto, Toronto, Ontario, Canada; Sunnybrook Research Institute, Sunnybrook Health Sciences Centre, University of Toronto, Toronto, Ontario, Canada; Divisions of Urogynecology and Minimally Invasive Gynecologic Surgery, Department of Obstetrics and Gynecology, Sunnybrook Health Sciences Centre, University of Toronto, Toronto, Ontario, Canada; Division of Maternal Fetal Medicine, Department of Obstetrics and Gynecology, Sinai Health System, University of Toronto, Toronto, Ontario, Canada; Division of Maternal Fetal Medicine, Department of Obstetrics and Gynecology, Sunnybrook Health Sciences Centre, University of Toronto, Toronto, Ontario, Canada; Division of Orthopedics & Trauma Surgery, Department of Surgery, Sunnybrook Health Sciences Centre, University of Toronto, Toronto, Ontario, Canada

## Abstract

**Introduction:** The Omicron variant of the SARS-CoV-2 virus is described as more contagious than previous variants. We sought to assess risk to healthcare workers (HCWs) caring for patients with COVID-19 in surgical/obstetrical settings, and the perception of risk amongst this group.

**Methods:** From January to April, 2022, reverse transcription polymerase chain reaction was used to detect the presence of SARS-CoV-2 viral RNA in patient, environmental (floor, equipment, passive air) samples, and HCW’s masks (inside surface) during urgent surgery or obstetrical delivery for patients with SARS-CoV-2 infection. The primary outcome was the proportion of HCWs’ masks testing positive. Results were compared with our previous cross-sectional study involving obstetrical/surgical patients with earlier variants (2020/21). HCWs completed a risk perception electronic questionnaire.

**Results:** 11 patients were included: 3 vaginal births and 8 surgeries. 5/108 samples (5%) tested positive (SARS-CoV-2 Omicron) viral RNA: 2/5 endotracheal tubes, 1/22 floor samples, 1/4 patient masks and 1 nasal probe. No samples from the HCWs masks (0/35), surgical equipment (0/10) and air samples (0/11) tested positive. No significant differences were found between the Omicron and 2020/21 patient groups’ positivity rates (Mann-Whitney U test, p = 0.838) or the level of viral load from the nasopharyngeal swabs (p = 0.405). Nurses had a higher risk perception than physicians (p = 0.038).

**Conclusion:** No significant difference in contamination rates were found between SARS-CoV-2 Omicron BA.1 and previous variants in surgical/obstetrical settings. This is reassuring as no HCW mask was positive and no HCW tested positive for COVID-19 post-exposure.

## Background

The Omicron variant of the SARS-CoV-2 virus has been reported to be more contagious than the previously encountered variants^1,2^. It contains numerous mutations in the spike protein allowing the virus to evade neutralizing antibodies ^1–4^. As a result, following 2 doses, vaccine efficacy at preventing infection was shown to be decreased to 70%, which is notably lower than has been reported against other variants^5^. Due to its high rate of transmission and reduced effectiveness of existing vaccines at preventing infection, the risk that the Omicron variant of the SARS-CoV-2 virus poses to healthcare workers (HCWs) in close contact with infected individuals is a topic of concern.

Surgery and obstetrics involve close and prolonged contact with patients. Transmission of the SARS-CoV-2 virus occurs primarily through respiratory droplets, aerosols and contact with contaminated surfaces^6,7^. However, the risk of HCWs contracting Omicron variant of the SARS-CoV-2 through respiratory droplets in an obstetrical or surgical setting is unclear. Besides the respiratory tract, the SARS-CoV-2 virus has also been shown to be present in the GI tract, amniotic fluid, peritoneal fluid and vaginal fluid^8–14^. For surgeries/procedures that involve these structures or substances, the risk of transmission through aerosolization of viral particles from the surgical site is unclear. Transmission can also occur by coming into contact with contaminated surfaces^6,7^. It is unclear whether HCWs involved in a surgery or an obstetrical delivery are at risk of being infected with the Omicron variant of the SARS-CoV-2 virus in such a manner.

HCWs’ perception of safety in their work environments has been investigated, particularly during the early stages of the pandemic^15–19^. Previous studies conducted around the world have indicated that personal protective equipment (PPE) shortage, insufficient protection provided by the PPE and fear of infection have all contributed to HCWs feeling unsafe at their workplaces^15–19^. However, whether these sentiments are shared by HCWs in Canada during more recent waves of the pandemic is not known.

In general, the risk to health care workers caring for patients infected with the Omicron variant in surgical and obstetrical settings is unclear. The aim of this study is to determine the risk associated with exposure to Omicron variant of the SARS-CoV-2 virus in the operating room or obstetrical delivery setting and whether this risk is higher in comparison to previous variants.

Further, we also seek to explore HCWs’ perception of risk at their workplace and their attitudes towards the PPE provided.

## Methods

### Study Design

A cross-sectional study was conducted from January 2022 to April 2022. Patients with a nasopharyngeal (NP) or mid-turbinate swab positive for SARS-CoV-2 by real-time reverse transcription polymerase chain reaction (RT-PCR), in need of urgent surgery or obstetric delivery at Sunnybrook Health Sciences Centre, were prospectively identified by the surgical or obstetric clinical teams. Urgent surgery, as per Sunnybrook’s operating room policies, was defined as 1A, 1B, 1C and 1D: surgery required within 2 hours, 2–8 hours, 8–48 hours and 2–7 days, respectively, to avoid harm to patients^20^. The Strengthening the Reporting of Observational Studies in Epidemiology (STROBE) checklist was used to report this study^21^.

### 2020-21 Study group^14^

A cross-sectional study was conducted from November 2020 to May 2021 at 2 tertiary academic Toronto hospitals (Sunnybrook Health Sciences Centre and Sinai Health System), during urgent surgeries or obstetric deliveries for 32 patients with SARS-CoV-2 infection. The presence of SARS-CoV-2 viral RNA in patient, environmental and air samples was identified by real-time reverse transcription polymerase chain reaction (RT-PCR).

Results of the 2020-21 Study group were compared with the current study 2022 Omicron group.

### Setting (2022)

Patients were recruited from Sunnybrook Health Sciences Centre, a level 3 obstetrical unit and a tertiary care regional trauma and burn centre.

Standard hospital procedures and PPE worn by health care workers attending patients with SARS-CoV-2 infection included the following: disposable protective head covering, mask (either N95 mask or American Society for Testing and Materials [ASTM] level 3 surgical mask), face shield or eye protection, impermeable gown, gloves, and shoe or boot covers. When possible, contaminated PPE was removed in the adjacent anteroom of the operating room. During intubation and extubation, only the anesthesia team (with N95 masks) remained in the room.

N95 mask fit testing was a requirement of hospital health care workers in the surgical and obstetric clinical areas. If able, patients wore ASTM level 3 ear loop masks. All the operating rooms (including those in the birthing area) were equipped with 20 air exchanges per hour.

### Participants

Patients were included if they were at least 18 years of age; were within 30 days of a positive nasopharyngeal swab for SARS-CoV-2 (either asymptomatic or symptomatic for COVID-19^22^) or were beyond 30 days from an initial positive nasopharyngeal swab for SARS-CoV-2 and still had symptoms of COVID-19, and required obstetrical delivery or urgent surgery.

Health care workers included any consenting health care workers present in the operating or delivery room, caring for the patient.

### Data Sources

Information about the patient’s history and presenting problem was extracted from the patient electronic medical records (EMR).

### Study Procedures

Study procedures are described in detail in Appendix 1. Patient sampling for abdominal surgical cases included peritoneal cavity fluid (male or female). Patient samples for obstetrical cases included vaginal fluid and swabs of the membranous placenta.

Equipment and environmental samples included swabs of the room floor (within 1 m from the surgical site, and 2 m away from the surgical site^23^), swab of equipment (e.g., endotracheal tube and surgical instruments), and swab of the inside of the surgical mask worn by health care workers^24–26^

Passive air sampling was performed using an open Petri dish to collect any viral particles settling by gravity in the dish (within 1–2 m of the patient, 1 m off the floor)^25,27–30^.

### Laboratory processes

All samples were processed at Sunnybrook Health Sciences Centre and the laboratory staff were blinded to the source of the sample.

Virus detection was performed by real-time RT-PCR using a multitarget assay^31^. The viral RNA loads from samples were extracted using the EasyMag Platform (bioMérieux, France) according to the manufacturer’s instructions. Detection of the SARS-CoV-2 viral RNA was performed using the Luna Universal Probe One-Step RT-qPCR Kit (New England Biolabs, Canada) with the primers and probe for the E-gene, and the thermocycling conditions that have been described by Corman and colleagues^32^ on the Rotor-Gene Q platform (Qiagen, Germany).

The cycle threshold (Ct) value of the assay was used as an estimate of the viral load and was obtained for all samples where possible, including values from the patient’s initial diagnostic swab. Samples were considered negative if the Ct value was 40 or greater.

### Survey of HCWs

HCWs who consented to have their mask swabbed and tested were contacted via email once within 6 weeks of the procedure and invited to participate in an electronic survey via LimeSurvey (Appendix 2). This 7-10 question survey assessed the HCWs’ experience of caring for a patient with a NP swab positive for COVID-19. HCWs were asked to indicate how safe they felt with the PPE provided on a Likert scale of 1(not safe at all) to 10 (very safe). The survey also included questions about whether any COVID-19 symptoms were experienced after the patient encounter, any COVID-19 testing they undertook after the procedure, and their vaccination status. The same survey was distributed in 2021 to HCWs who participated in an earlier study^14^, caring for NP swab positive patients between November 2020 to May 2021 approximately 4-6 months after their exposure.

### Statistics

Not all patients had similar data as there were different clinical scenarios. Descriptive statistics were generated using Microsoft Excel (Version 16.62) for: age of the patients, Ct values of the nasopharyngeal swabs and the survey results. Further, the Mann-Whitney U test was performed using R (Version 4.2.1, 2021) to: compare NP swab Ct values of patients with a positive RT-PCR study sample and patients with no positive study sample, and NP swab Ct values of patients with the Omicron variant of the SARS-CoV-2 virus and other patients with other previous variants (from the 2020-21 Study group)^14^. The HCW survey results from different groups of HCWs (e.g. physicians, nurses,) were also compared using the Mann-Whitney U test. The 2-sample test for equality of proportions was used to compare the sample positivity rate in the current study group and the 2020-21 study group. Additionally, the Shapiro-Wilk test was used to assess for normality at all times before using the Mann Whitney U tests.

### Outcomes

The primary outcome was the proportion of health care workers’ mask samples with positive SARS-CoV-2 RNA RT-PCR results. Other outcomes included the rate of SARS-CoV-2 RNA PCR-positive samples from the surgical site (relevant patient samples), surgical equipment, floor, and ambient air of the operating room or birthing room. Additionally, as described in the “Laboratory processes” section, the Ct value was obtained for all positive samples where possible. For the HCW survey, main outcomes of interest included number of HCWs who contracted COVID-19 as a result of the exposure in the operating room or birthing room, and how safe they felt with the PPE provided.

### Ethics Approval

The Sunnybrook Research Ethics Board granted ethics approval (#1676).

Patient samples (peritoneal fluid, vaginal, or placental swabs) were collected in patients providing informed consent. Mask sampling of attending health care workers was performed with health care workers’ consent. Patient or health care worker consent was not required for sampling from the air, floor, or surgical instruments. Consenting health care workers agreed to follow up with hospital occupational health departments if SARS-CoV-2 RNA was detected on the inside surface of their mask, and also agreed to participate in an online survey emailed to them after the patient encounter.

## Results

### Description of the cohort

Eleven patients at Sunnybrook Health Sciences Centre were recruited for the study, five were male and six were female (mean age 43.8 years, standard deviation 14.9). Three of the patients had vaginal deliveries, while the other eight underwent urgent surgery. A nasopharyngeal swab testing for SARS-CoV-2 was performed on all patients in this study at a median of 3 days before their procedure (mean 4.63, range 1-16). Two patients underwent a repeat nasopharyngeal swab on the day of their procedure. The vaccination status for 10/11 of the patients was known. Three of the patients received three doses of the COVID-19 vaccination; 5 patients received two doses of the COVID-19 vaccinations and 2 were unvaccinated.

### Detection of SARS-CoV-2 in surgical and environmental samples

A total of 108 samples were collected (Table 1). Five of 108 samples were duplicates (endotracheal tube sampled using two different methods). All samples were tested for the presence of the SARS-CoV-2 virus and five out of the 103 samples (5%) tested positive: 2 out of 5 endotracheal tubes (2 urgent surgery cases), 1 out of 22 floor samples (1 neurosurgery case, sample from at least 2 metres away from the bed) and 1 nasal temperature probe sample (Burn Surgery case). A swab taken from the inside of 1 patient’s mask was positive (vaginal birth). None of the samples from the HCWs masks (0/35), surgical equipment (0/10) and Petri dishes (0/11) tested positive. In the 2020-21 study group, 20/332 samples (6%) had tested positive for SARS-CoV-2 RNA. No significant differences were found between the sample positivity rates of this Omicron study group and the 2020-21 study group of earlier SARS-CoV-2 variants/subtypes (2 sample test for equality of proportions, p-value = 0.838)

**Table 1:**
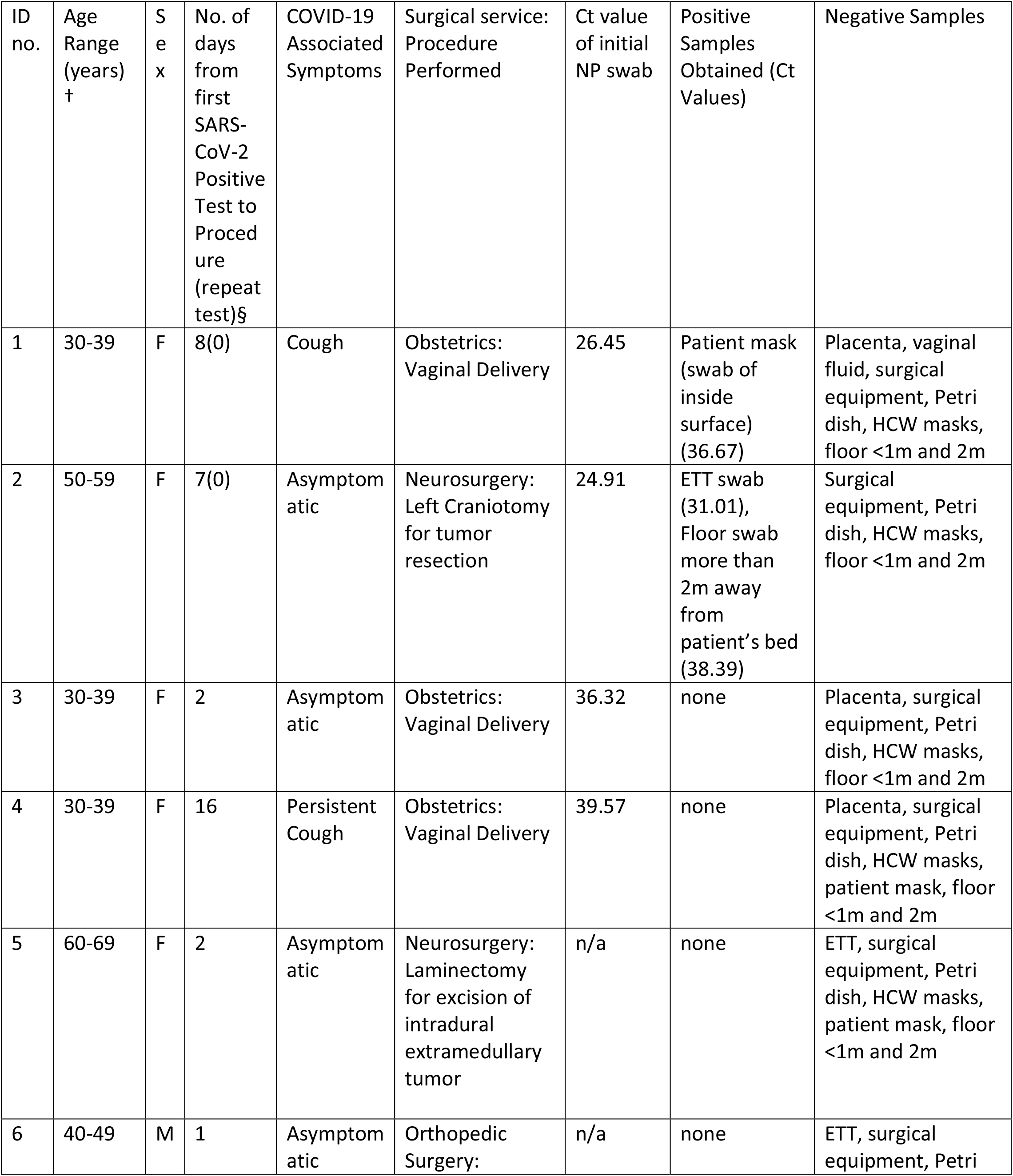
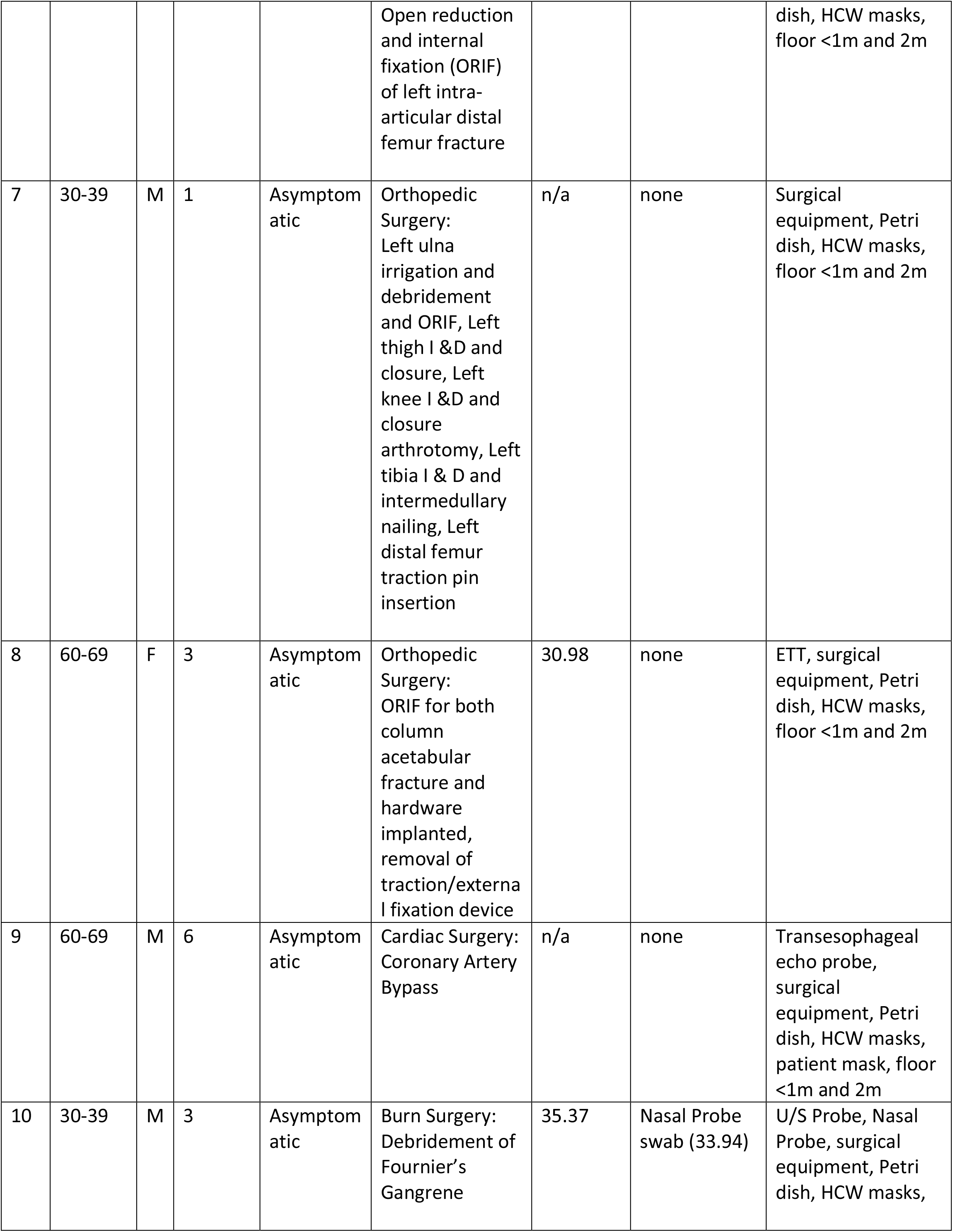
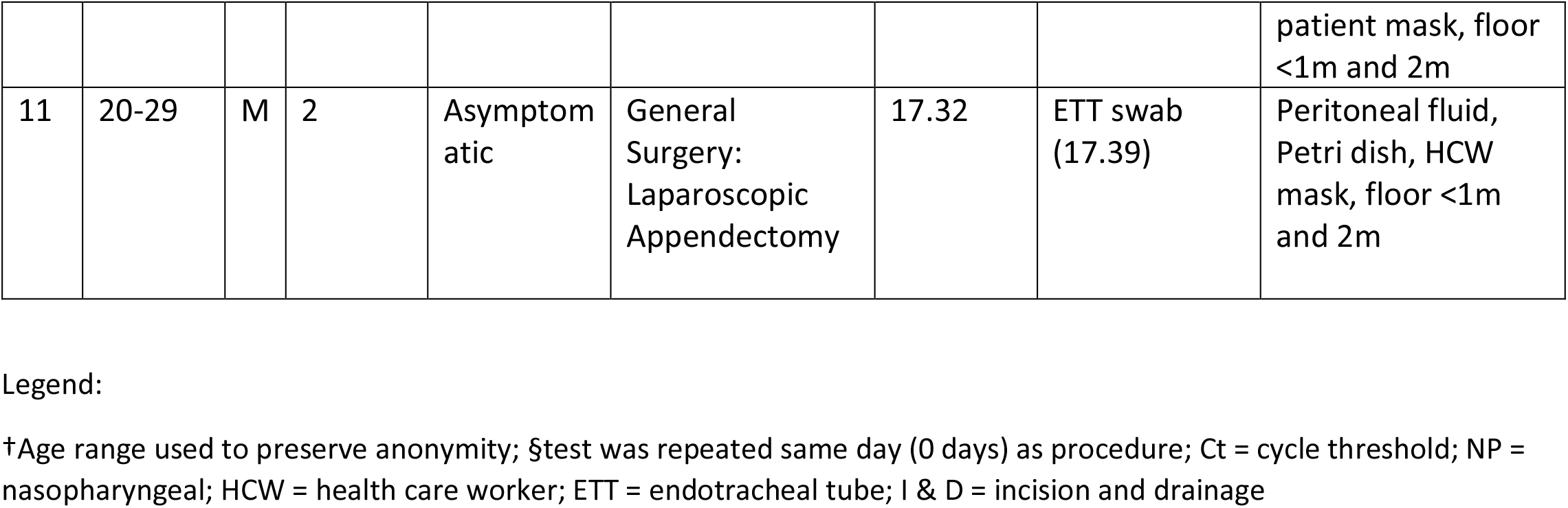
Clinical Case Details and Results of Samples Obtained.

### Viral Load in Patient Samples

During our study period of January to April 2022, the SARS-CoV-2 variant isolated in the Sunnybrook Microbiology Lab was only Omicron BA.1. For 7/11 patients in this study, Ct values were obtained from the nasopharyngeal swabs that were collected (mean = 30.13, range 17.32 – 39.57). Comparing the NP swab Ct values of the 4 patients with any positive study sample (of surgical equipment, environment) and the 3 patients with no positive study sample, there were no significant differences found between the two groups (Mann-Whitney U test, p = 0.114).

NP swab Ct values were also collected for the 2020-21 study group, consisting of 32 patients with previous variants of the SARS-CoV-2 virus (mean = 26.95, range 11.86 – 37.25)^14^. When compared with NP swab Ct values of patients with the Omicron variant, no significant difference was found between these two groups of patients (Mann-Whitney U test, p = 0.405).

### HCW Survey

A total of 45 responses were recorded for the survey for a total of 70 patient case encounters (with a HCW potentially having more than one patient encounter): 13 nurses, 29 physicians (surgeons, anesthetists, or their house staff) and 3 respiratory technician/anesthesia assistants. Of the 45 HCWs, 32 are from the time period of Nov 2020-May 2021 (the 2020-21 group) and 13 are from the period of January-April 2022 (the 2022 Omicron group). The survey response rate was 37% (29/78) for the 2020-21 group and 48% (13/27) for the 2022 Omicron group.

Three of 45 HCWs arranged for COVID-19 testing within 2 weeks after the clinical exposure: 1 was worried about the exposure (2020-21 group), 1 had symptoms (2020-21 group) and 1 was asymptomatic but arranged for testing due to upcoming planned travel (2022 Omicron group). All 3 tests were reported as negative. Zero of 45 HCWs reported being ill with COVID-19 within 2 weeks after clinical exposure. Two of 45 HCWs reported not being vaccinated at the time of clinical exposure (2020-21 group); 8/45 had 1 vaccination dose and all others (35/45) had at least 2 doses of COVID-19 vaccine. HCWs were asked to indicate on a Likert scale of 1 (not safe at all) to 10 (very safe) how safe they felt with the PPE provided. The average score was 8.066 (range 3-10). There was a significant difference between the scores given by physicians (mean 8.69 +/−1.54) and nurses (mean 6.92 +/−2.43), Mann U Whitney p-value = 0.0377, Table 2 and Figure 1. Sixty-five percent (19/29) of physicians indicated 9 or 10 on the Likert scale for how safe they felt with the PPE they were provided compared to 31% (4/13) of nurses who responded 9 or 10.

**Figure 2:**
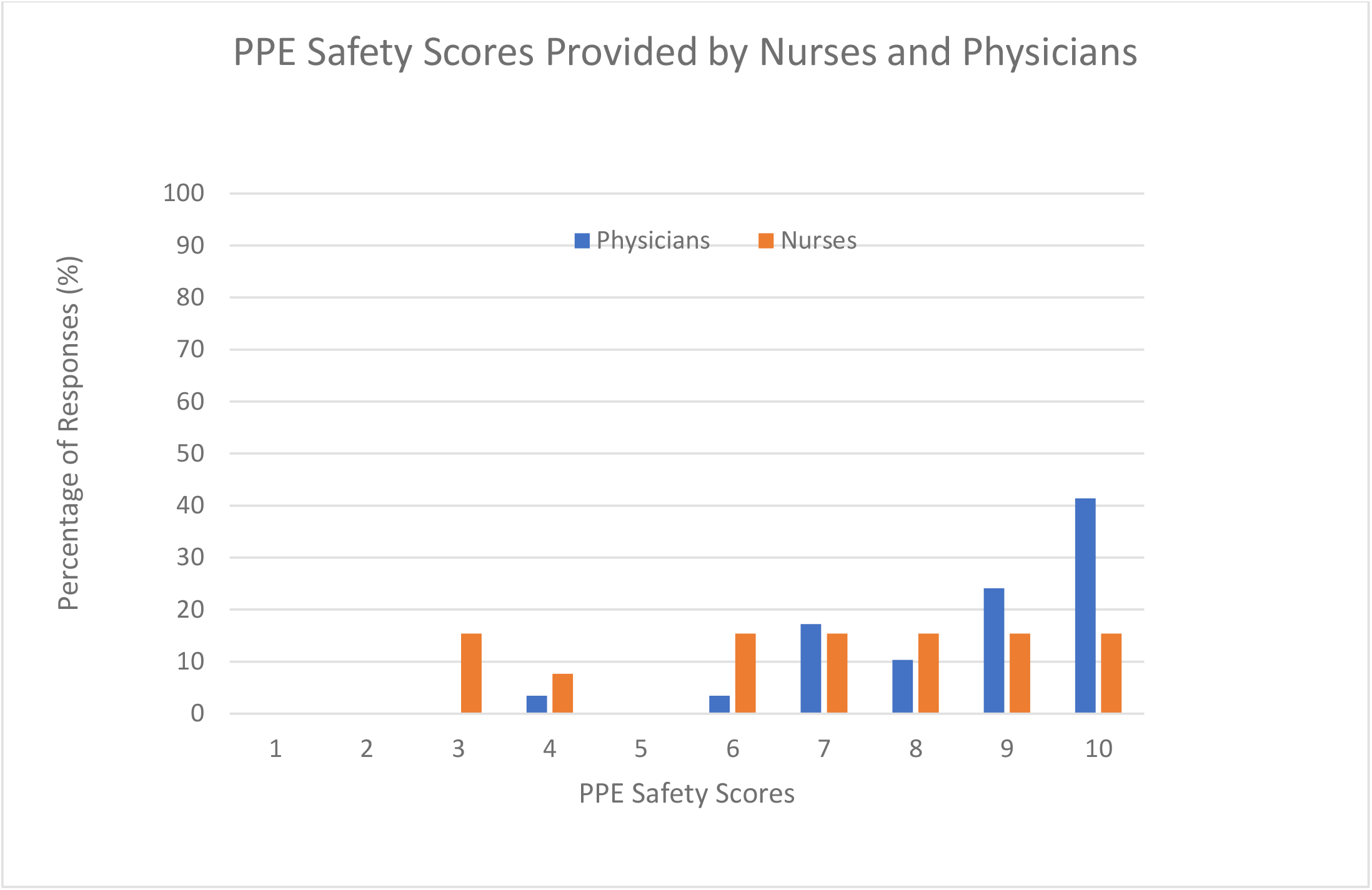
Personal Protective Equipment (PPE) safety scores.

**Table 2:** Health care worker Survey after the exposure to a patient infected with SARS-CoV-2.

| HCW role | No. of Responses | No. of HCWs who arranged to be tested after the exposure | No. of HCWs who experienced symptoms after the exposure | No. of HCWs who became sick with COVID-19 after the exposure | Mean score given for how safe HCWs felt with the PPE provided (± SD) [Likert scale: 1 = felt not safe at all, 10 = felt very safe] |
| --- | --- | --- | --- | --- | --- |
| Anesthesia Assistant | 2 | 0 | 0 | 0 | 5.50±3.54 |
| Respiratory Technician | 1 | 0 | 0 | 0 | 10<sup>†</sup> |
| Nurse | 13 | 1 | 1 | 0 | 6.92±2.43 |
| Physician | 29 | 2 | 0 | 0 | 8.69±1.54<sup>‡</sup> |

HCW = health care worker; PPE = personal protective equipment
<sup>†</sup>There was only one response for this category, mean not calculated

## Discussion

In the era of COVID-19, the safety of HCWs involved in surgery and obstetrics is paramount due to the prolonged and close contact they have with potentially infected patients. We found no difference in contamination rate in the operating room and birthing room settings between this study’s Omicron BA.1 group of 11 patients and our previous study group of 32 patients from 2021-21, representing a cohort infected with previous variants of concern.

With this study, none (0/35) of the swabs collected from the inside surface of HCW masks tested positive for the SARS-CoV-2 viral RNA. This is similar to the results from our 2020/2021 study group, which had also found no evidence of viral RNA on the inside surface of HCWs’ masks^14^. Brandner et. al also found no contamination of masks in HCWs that attended autopsies: they assessed contamination of PPE during autopsies of COVID-19 patients and reported that samples from gloves, aprons and tops of shoes worn by HCWs tested positive for the SARS-CoV-2 viral RNA^33^. Infectious virus was isolated from 21% of the RNA-positive glove samples (3/11 full autopsies)^33^. Similar to surgery/obstetrics, autopsies may require close and prolonged contact with body fluids and secretions and consequently the aforementioned findings of PPE contamination may also be applicable to surgical or obstetrical settings. Thus, it is important to consider that while the samples from the masks of HCWs had tested negative, a risk of transmission still remains.

Out of the 108 samples that were tested for this study, five study samples tested positive (5%). Among the samples that tested positive are 2 endotracheal tubes (ETT) samples and 1 nasal temperature probe sample. The positivity of the ETT and nasal probe samples can be attributed to the presence of SARS-CoV-2 in the respiratory tract^7,22,34,35^. The inside surface of one patient mask tested positive (obstetrical patient).The positive floor swab is consistent with studies conducted in the ICU that have reported contamination of the air and surfaces within about 4m from patients infected with SARS-CoV-2^36^. Further, these findings are comparable to that of the 2020/2021 study group where 8 of 12 ETT samples and 5 out of 60 floor samples tested positive^14^.

Passive air sampling in this current study using the petri dish did not yield any positive results. A study by Schoen et. al investigating the presence of viral RNA in the air in the labor room with un-vaccinated patients who had tested positive for SARS-CoV-2 had also failed to detect the viral RNA in the air^37^. However, in our previous study, 1of 7 passive air samples in the operating room had tested positive for the SARS-CoV-2 RNA^14^. Also, de Man et. al that found 4 in 17 air samples of the Intensive Care Unit (ICU) to be positive for SARS-CoV-2^38^.

None of the surgical equipment samples for this study had tested positive (other than ETT, nasal probe), whereas, in our previous study, 2 of 11 samples from surgical equipment (scissors, clamps) had tested positive^14^. These low sample positivity rates align with findings of a study by Fabbri et.al that indicate that the transmission risk during surgery, specifically through aerosolization from the surgical field, is low^39^.

Generally, the contamination rates observed for the Omicron variant (5%) in this study and the previous variants (6%) from our previous study have been quite similar. There was no significant difference in the NP swab Ct values between these two groups which would indicate that the viral load is not higher with one group compared to the other, although this may also be due to the small sample sizes. Glirnet et al also found no significant difference in contamination rates between the original, Alpha and Omicron strains of the SARS-CoV-2 virus in a COVID-19 patient ward setting: environmental samples from a hospital isolation ward and a quarantine hotel during February 2021 for the Alpha and original variants and in January 2022 for the Omicron variant^40^. Although we did find in our previous study that higher viral load (lower NP Ct value) was associated with a higher risk of contamination^14^, we did not demonstrate a similar finding with this Omicron study, which may have been due to the smaller number of patients studied.

In our study with 3/45 HCWs arranging for testing after the exposure, it can be inferred these HCWs generally perceived their work environment to be safe. Further, most of the survey respondents in both the 2020/21 group and the 2022 Omicron group indicated that they felt safe with the PPE they were provided. This contrasts with the results of the previous studies reporting low HCW satisfaction with PPE provided and safety at work^15–19^. In a study that was conducted in Qatar from June 2020 to July 2020, Ismail et. al reported that only 45.6% of HCWs surveyed were satisfied or highly satisfied with the quality of the PPE provided to them^16^.

Similarly, a study by Aloweni et. al conducted in Singapore from July 2020 to September 2020 also found that only 13.7% of the participating HCW were ‘highly confident’ of the protection provided by the PPE^17^. However, the measures used to evaluate “feeling safe” or “satisfaction with” or “being confident” with the PPE used in these studies were different, and it is also important to keep in mind that these studies took place in different countries during a different time period when PPE shortage could have been possibly a more pertinent issue^15–19^.

Interestingly, in our HCW survey we found a significant difference between nurses and physicians in terms of how safe they felt with the PPE they were provided. Nurses had responded with lower scores compared to physicians with 54% of nurses compared to 25% of physicians scoring 7 or lower to indicate how safe they felt with the PPE provided. It is possible that being involved in caring for the patient for longer time periods and spending more time in the operating room and birthing room environment may have all contributed to these lower scores.

## Limitations

This study has limitations. Firstly, the study size is small; therefore, it is possible that the results cannot be generalized to the rest of the population with the Omicron variant of the SARS-CoV-2 virus. Further, due to limitations of testing procedure and equipment, it is possible that the presence of the SARS-CoV-2 virus may not have been detected in certain samples. It is also recognized that capturing positive air samples may be challenging^41^. In the future, studies can be conducted to further quantify the risk to HCWs caring for patients in a surgical or obstetrical setting by looking for and documenting the presence of infection amongst HCW post-exposure. The HCW survey response may have been higher with frequent email reminders.

## Conclusion

Despite the Omicron variant being described as more contagious, there does not appear to be an increased risk of SARS-COV-2 viral RNA contamination in the operating room or birthing room environment with the Omicron BA.1 variant compared to earlier subtypes and variants of concern.

## Data Availability

All data produced in the present study are available upon reasonable request to the authors.

## APPENDIX 1: Supplement for Study Sample Collection

### GENERAL DETAILS

Each dental pledget was pre-moistened with up to 3cc of sterile UTM from a newly opened unused 15cc sterile Falcon tube containing 3 cc of UTM, universal transport medium (UTM®, Copan Diagnostics, CA, USA; https://www.copanusa.com/sample-collection-transport-processing/utm-viral-transport/

Hand hygiene and glove changes were performed between the collection of every sample and the outside surface of each container was wiped off with a CaviWipes^™^ towelette (https://www.metrex.com/en-ca/caviwipes) and then each sample was stored in a separate new biohazard marked ziploc plastic bag and placed in the fridge at 4° Celsius within 20 minutes. All collected samples were processed at the Sunnybrook Microbiology Lab (stored in the interim at −20° and then −80° Celsius).

Samples were analyzed by RT-PCR as previously described (Vermeiren C, Marchand-Senecal X, Sheldrake E, et al. Comparison of Copan Eswab and FLOQswab for COVID-19 PCR diagnosis: working around a supply shortage. J Clin Microbiol 2020.)

All the operating rooms (including those in the Birthing area) have 20 air exchanges an hour.

Cycle threshold values (the number of cycles required for the fluorescent signal to cross the threshold in RT-PCR) quantified viral load, with lower values indicative of a higher viral load.

### PATIENT SAMPLES

#### 1) PERITONEAL FLUID SAMPLE

Taken by a member of the surgical team: 5-10 cc of peritoneal fluid (if present upon entering the cavity), or, if no fluid seen, 10cc of sterile saline was placed in the peritoneal cavity with a 10cc sterile syringe and then whatever volume of fluid aspirated back into the syringe was placed in an 80cc sterile plastic container.

#### 2) VAGINA

Taken by a member of the obstetrical team: a vaginal speculum was placed in the vagina before delivery (typically after informed consent and well before active labor) and up to 5 cc of pooled vaginal fluid was aspirated with a sterile 10cc syringe. If no vaginal fluid was seen, 10cc of sterile saline was placed in the vagina with a 10cc sterile syringe and then whatever volume of fluid that was aspirated back into the syringe was placed in an 80cc sterile plastic container.

##### 3) PLACENTA

Taken by a member of the obstetrical/research team at the end of the case with a flocked swab (iClean, HCY, Shenzhen, China; https://www.chenyanglobal.com/oropharyngeal-nylon-flocked-swab-product/) which was used to wipe the surface of the membranous placenta and the swab was placed immediately in a 15cc sterile plastic Falcon tube containing 3cc of UTM.

#### ENVIRONMENTAL SAMPLES

#### 4) FLOOR

Taken by a member of the research team at the end of the case: a sterile dental pledget (3/8” x 1.5” cylindrical sponge, SDP Inc. Montreal, Canada; https://www.sdpmedical.com/en/cylindrical-sponges) was pre-moistened with sterile universal transport media and the floor was swabbed in a location as close to the patient as permits and also at least 2 metres away. The swabbing was performed in a standardized fashion over a 30×30cm area with 2 perpendicular “S” swipes. The pledget was placed immediately in a 15cc sterile plastic Falcon tube containing 3cc of UTM.

#### 5) ENDOTRACHEAL TUBE (ETT)

Taken by a member of the research team at the end of the case after the patient was extubated: a sterile dental pledget or a flocked swab was pre-moistened with UTM and used to wipe the length of the distal half of the ETT and the pledget/swab placed immediately in a 15cc sterile plastic Falcon tube containing 3cc of UTM.

#### 6) SURGICAL INSTRUMENTS / EQUIPMENT

Taken by a member of the research team at the end of the case: a sterile dental pledget was pre-moistened with sterile UTM and used to wipe the part of the instrument or equipment that was in direct contact with the patient’s surgical site. The pledget then placed immediately in a 15cc sterile plastic Falcon tube containing 3cc of UTM.

#### 7) PASSIVE AIR SAMPLE

3cc of sterile UTM was placed in a sterile 90mm Petri dish which was placed open by a member of the research team at the beginning of the case within 1-2 metres of the patient and in a location that would not interfere with patient care, on a Mayo stand about 1 metre high from the floor. The Petri dish was retrieved at the end of the case and the UTM in it transferred to a sterile Falcon tube.

#### 8) MASK SAMPLE

Masks were swabbed on their inside surface by a member of the research team with a sterile dental pledget that was pre-moistened with UTM – wiping over the inside of the mask twice in the area (up to 10×10cm) that would have been in contact with the nose and mouth. The pledget then placed immediately in a 15cc sterile Falcon tube containing 3cc of UTM.

## APPENDIX 2: Health care worker Questionnaire (via *LimeSurvey*)

**Q1**: How many times did you participate in this COVID-19 study and leave your mask to be tested with a study team member?

a) 1 b) 2 c) 3 d) 4 e) 5

**Q2**: Please indicate your role

a) Nurse (including scrub nurse, circulating nurse,student) d) Respiratory Technician

b) Physician - anesthesia team member (including staff, house staff, student) e) Prefer not to say

c) Physician - surgical team member (including staff, house staff, student) f) Other:________

**Q3**: Within 2 weeks after you worked with a COVID-19 positive patient in the OR (or birthing unit), did you arrange to be tested for COVID-19?

a) Yes b) No (go to Q6)

**Q4**: Did you arrange to be tested because you were worried about the exposure?

a) Yes c) Not sure

b) No Free text option:________

**Q5**: Within 2 weeks after you worked with a COVID-19 patient in the OR (or birthing unit), did your COVID-19 swab/test result return positive for COVID-19

a) Yes c) Not sure

b) No Free text option:________

**Q6**: Within 2 weeks after you worked with a COVID-19 positive patient in the OR (or birthing unit), did you develop COVID-19 symptoms?

a) Yes b) No Free text option:________

**Q7**: Within 2 weeks after you worked with a COVID-19 positive patient in the OR (or birthing unit), did you become sick with COVID-19?

a) Yes b) No (go to Q9) Free text option:________

**Q8**: If you tested positive for COVID-19 or were sick with COVID-19 within 2 weeks after you worked with a COVID-19 patient in this study, do you think you became COVID-19 positive as a result of this exposure?

a) Yes d) I don’t know

b) No - I think I became sick from a different exposure (a different patient) Free text option:

c) No - I think I became sick from a different exposure (not from a patient at my workplace)

**Q9**: Were you vaccinated at the time of study participation?

a) Yes, I had at least 2 doses of the COVID vaccine c) I was not vaccinated at all at the time of exposure

b) I had only 1 dose of COVID vaccine at the time of the exposure Free text option:

**Q10**: I felt safe with the PPE provided for the case:

Likert scale

response: 1

(not safe at all) 2 3 4 5 6 7 8 9 10

(very safe)

Free text option for any comments:________

